# Proactive vs. reactive COVID-19 screening in schools: lessons from experimental protocols in France during the Delta and Omicron waves

**DOI:** 10.1101/2025.01.17.25320676

**Authors:** Elisabetta Colosi, Bruno Lina, Christelle Elias, Philippe Vanhems, Vittoria Colizza

**Affiliations:** Sorbonne Université, INSERM, Pierre Louis Institute of Epidemiology and Public Health, Paris, France; National Reference Center for Respiratory Viruses, Department of Virology, Infective Agents Institute, Croix-Rousse Hospital, Hospices Civils de Lyon, Lyon, France; Centre International de Recherche en Infectiologie (CIRI), Virpath Laboratory, INSERM U1111, CNRS—UMR 5308, École Normale Supérieure de Lyon, Université Claude Bernard Lyon, Lyon University, Lyon, France; Service Hygiène, Epidémiologie, Infectiovigilance et Prévention, Hospices Civils de Lyon, Lyon, France; Centre International de Recherche en Infectiologie, Institut National de la Santé et de la Recherche Médicale U1111, Centre National de la Recherche Scientifique Unité Mixte de Recherche 5308, École Nationale Supérieure de Lyon, Université Claude Bernard Lyon 1, Lyon, France; Department of Biology, Georgetown University, Washington, District of Columbia, USA

## Abstract

The COVID-19 pandemic prompted diverse policies to manage safety in schools, balancing infection control with educational continuity. This study assessed the impact of an experimental weekly screening protocol compared to nationally implemented reactive strategies in 25 primary schools in the Auvergne-Rhône-Alpes region of France during the Delta (November–December 2021) and Omicron (January–February 2022) waves. We used an agent-based model for SARS-CoV-2 transmission in schools parameterized with empirical data characterizing school contact over time to estimate the contribution of school transmission on overall cases and evaluate the effectiveness of weekly screening in reducing within-school infections. We parametrized the model to reproduce the Delta and Omicron variants dominant in the study period, accounting for introductions from community surveillance data. We fitted the model to the observed prevalence in 18 schools selected for the analysis. School transmission was estimated to account for 67% (IQR 53-78) of student cases in Rhône and 67% (IQR 50-82) in Savoie during the Delta wave, and 52% (IQR 47-57) in Rhône during the Omicron wave. The experimental weekly screening protocol was estimated to reduce transmission in school by 40% (IQR 18 – 53%) during the Delta wave and by 37% (IQR 30-45) during the Omicron wave, compared to the reactive strategies applied in the same period in the rest of the country. Adherence rates exceeding 80% during the study were critical to the protocol’s success, contributing to an earlier and sustained decline in prevalence. Weekly screening proved a more structured and effective approach to controlling transmission, supporting its inclusion in future pandemic preparedness plans to ensure safer learning environments. This study underscores the importance of proactive interventions to address asymptomatic spread in schools, emphasizing their role in pandemic response strategies.

## INTRODUCTION

The COVID-19 pandemic has highlighted the diverse approaches adopted by governments worldwide to manage school safety while balancing the need for educational continuity^1^. Strategies have ranged from prolonged school closures^2^ and strict quarantines to the implementation of proactive testing and screening protocols. This variability reflects differences in public health priorities, resource availability^3^, and the perceived role of schools in viral transmission. These heterogeneous policies have underscored the complexity of managing outbreaks in educational settings, particularly given the significant social and developmental consequences of school disruptions^4,5^. France offers a unique case study of evolving school policies during the pandemic, transitioning from class closures to reactive screening strategies^6^ and later exploring systematic approaches^7^.

In September 2021, as schools reopened nationwide, France reinstated class closures in response to COVID-19 cases. This policy mandated quarantine for all students in a class upon the detection of a positive case, aiming to prevent further transmission among close contacts. Additional measures, such as mandatory mask-wearing and restricting interactions during meals or sport activities, were also implemented to limit exposure^8^. However, the start of the Delta wave in Fall 2021 led to an escalation of class closures, with nearly 2% of all classes disrupted by end of November 2021, and 73 out of 48,950 primary schools completely closed^9^. To mitigate the growing educational disruption, French authorities introduced a nationwide reactive screening strategy starting in week 49 of 2021 (December 6, 2021) as an alternative to class closure^6^. This strategy enabled in-person attendance by screening the entire class upon identifying a positive case, isolating only those who tested positive while allowing others to continue attending school.

Although reactive screening marked a shift toward minimizing educational disruptions, increasing evidence from modelling studies suggested that systematic screening on a weekly or semi-weekly basis would be more effective in controlling viral transmission^10–12^. Unlike reactive screening, which is triggered after a symptomatic case is detected, systematic screening proactively identifies asymptomatic infections, facilitating the timely isolation of positive cases and interruption of transmission chains. Reactive screening, on the other hand, risks missing undetected introductions and secondary infections, reducing its overall efficacy^13^. Based on these findings, French authorities piloted a weekly screening protocol as an experimental initiative in selected primary schools in the Auvergne-Rhône-Alpes region from week 47 to week 50 of 2021^7^. The objective of this protocol was to assess the effectiveness of weekly screening in reducing transmissions among students in real-world conditions by monitoring prevalence rates. Several participating schools opted to extend the weekly screening into January 2022, during the Omicron wave, providing further data for analysis.

In this study, we used an agent-based transmission model parameterized with empirical contact data from primary schools and fitted to the weekly screening results of the experimental protocol. We evaluated the impact of school transmission on viral circulation among primary school-aged children and assessed the effectiveness of weekly screening in reducing transmission during the Delta and Omicron waves. These results were compared to the nationally implemented reactive screening strategy. Our findings have important implications not only for managing SARS-CoV-2 in schools but also for informing strategies to maintain safe educational environments during future pandemics involving respiratory viruses. By providing evidence-based insights, this study contributes to the development of more effective and proactive public health policies for educational settings.

## METHODS

### Data collection under the experimental weekly screening

The experimental screening campaign targeted students aged 6-10 y.o. from 25 primary schools randomly selected in the departments of Isère, Puy-de-Dôme, Rhône, and Savoie within the Auvergne-Rhône-Alpes region. The campaign was conducted over two separate periods, divided by the Christmas holidays. Period 1 spanned weeks 47-50 of 2021 (November 22 to December 17, 2021) during the Delta wave, while Period 2 occurred during weeks 01-06, 2022 (January 3 to February 13, 2022) amidst the Omicron wave. In Period 1, over 99% of detected cases were identified as the Delta variant (week 43, 2021), with the peak incidence reaching 1,457 cases per 100,000 children aged 6-10 years (week 49, 2021). In contrast, Period 2 coincided with the emergence of the Omicron variant (96% BA.1 Omicron in week 02, 2022), which saw a peak incidence of 7,652 cases per 100,000 children in the same age group (week 03, 2022)^14^ (**Figure 1**). The primary objective of the experimental screening was to collect data on student prevalence and evaluate the effectiveness of weekly screening in preventing new cases.

**Figure 1.**
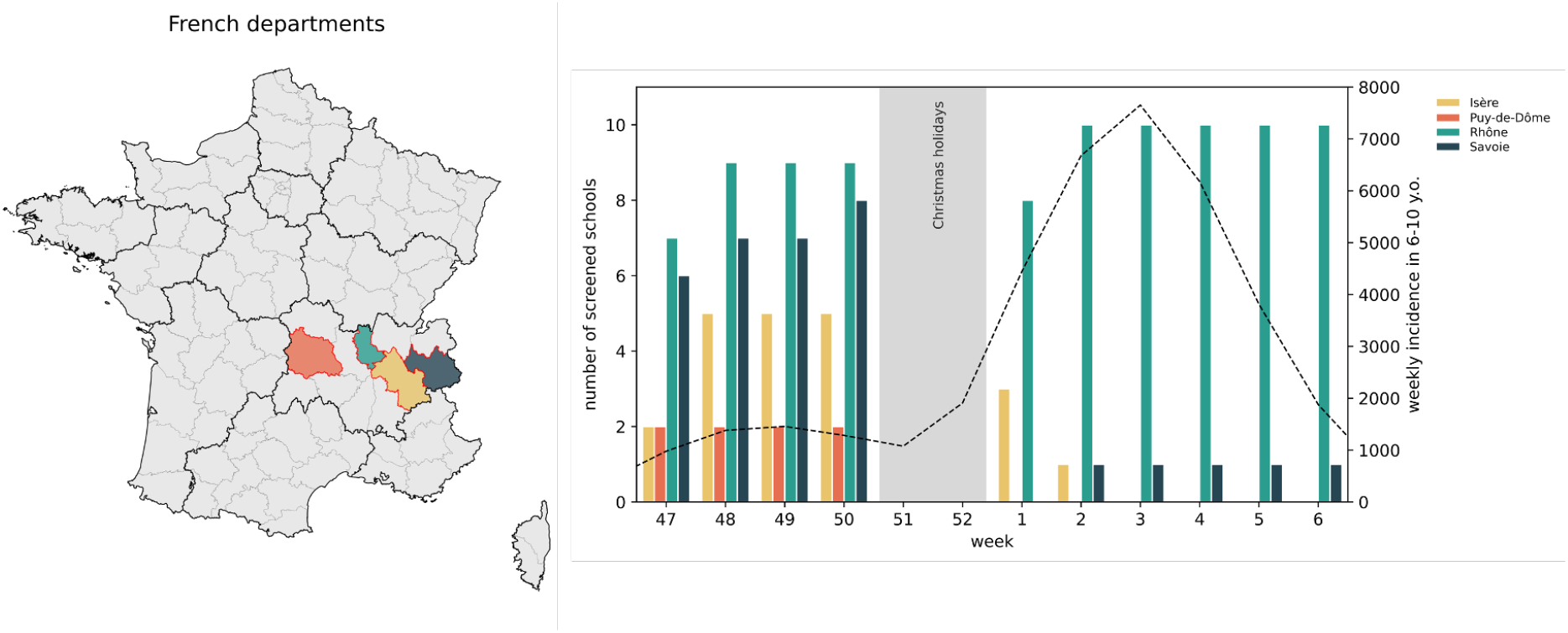
Selected departments, number of screened schools and epidemiological situation in weeks 47, 2021 - 06, 2022 (22/11/2021-13/02/2022). Number of screened schools for each department (solid bars) between week 47, 2021 and week 06, 2022. The four selected departments are: Isère, Puy-de-Dôme, Rhône and Savoie. The dashed line indicates the weekly incidence (cases per 100,000) from community surveillance in children aged 6-10 y.o., corresponding to primary school students. The Christmas holiday period (weeks 51–52, December 20, 2021–January 2, 2022) is shaded in light grey.

Over the experimental period (weeks 47, 2021 – week 06, 2022), weekly PCR tests on saliva samples were offered every Monday to students in participating primary schools. Participation was voluntary, requiring informed written consent from parents. Samples were collected at home, and the testing was conducted in collaboration between schools and medical laboratories. School principals provided class and participant lists to assigned laboratories, which managed the scheduling of tests and communicated results. Students who tested positive were required to isolate for seven days, with their status reported to local health authorities for follow-up. If more than three positive cases were detected within a single class, the entire class was closed for seven days.

For this study, we accessed anonymized, aggregated data for each participating school. These data included the number of students present on screening days, the number of tests conducted, and the number of positive cases. Only departments with at least five schools participating in weekly screenings (and at least 500 tested students per week) were included in the main analysis, in line with previous studies^10^. Sensitivity analyses were also performed, relaxing this criterion to include all schools participating in the campaign. Adherence to screening was computed as the ratio of tested students to the total number of students present on the screening day. Prevalence was determined as the ratio of positive tests to the total number of tests conducted.

### Nationwide school protocols

During the study period (week 45 of 2021 to week 06 of 2022), two changes in nationwide school protocols were implemented in France. Initially, the school protocol was relied on reactive class closures upon the detection of a case. Under this policy, the positive case and their classmates were required to isolate for seven days.

As case numbers began to rise during the Delta wave, authorities introduced a reactive screening protocol starting in week 49 of 2021^6^. Under this updated strategy, all students in the same class of a detected case were required to perform a RT-PCR or later-flow device (LFD) test on day 1 and day 7 following the detection of the index case. Only students with a negative test result were allowed to continue attending school in person, while positive cases were required to isolate for ten days. Additionally, the detection of three positive cases within a single class triggered a seven-day classroom closure.

When schools reopened in January 2022 following the Christmas holidays, a more stringent version of the reactive screening protocol was introduced in response to the emergence of the Omicron variant. Starting in week 01 of 2022, students in the same class as a detected case were required to complete an LFD or RT-PCR test on day 0, followed by two self-tests on days 2 and 4. Students with positive test results were required to isolate for seven days. Families were responsible for administering the tests, which could be performed at pharmacies, laboratories, or similar facilities outside the school. Beginning January 14, 2022, self-tests on day 0 were also accepted as an alternative to LFD or RT-PCR tests conducted at testing centers^15^. Unlike the previous protocol, no classroom closures were mandated under this strengthened strategy. However, local authorities retained the discretion to close classes in the event of a high number of cases.

### Ethical statement

Contact studies were approved by the Commission Nationale de l’Informatique et des Libertés (CNIL, the French national body responsible for ethics and privacy) and school authorities. Informed consent was obtained from participants or their parents if minors. No personal information of participants was associated with the RFID identifier. No ethical committee was required for this study as no individual data were collected in the study. All data retrieved during the screening campaign were anonymous and aggregated at the school level. Oral and written consent were obtained from parents ahead of the campaign.

### Transmission model

We designed a stochastic agent-based model of SARS-CoV-2 transmission in schools originally developed to study the Alpha wave in school settings^10^, parameterized with empirical contact data. The model used data on face-to-face proximity interactions, captured through RFID sensors with a 20-second time resolution, in a French school comprising 232 primary students and 10 teachers divided into 10 classes^16^. These sensor data were used to generate temporal contact networks, where nodes represent individuals (classified by class and student or teacher) and links represent empirically measured proximity contacts occurring at specific times. The contact networks revealed a strong community structure centred around classes, with students spending more time interacting within their own class than with students from other classes^10^. Additionally, students were observed to have longer interactions than teachers.

Transmission in the model occurs with a specific transmissibility rate (β/min) per contact per unit time between an infectious individual and a susceptible one who are in contact. The transmission rate ? was fitted to reproduce the observed student prevalence estimated from the experimental weekly protocol data (see Inference framework subsection). Infection progression includes prodromic transmission, followed by clinical or subclinical disease stages, informed from empirical distributions^17–19^. We also allowed individuals to enter the transient phase (*R*_+_) following the clinical or subclinical phase, where they are no longer infectious but can still test positive through a PCR test. The model was parameterized with age-specific estimates of susceptibility, transmissibility, probability of developing symptoms, and probability to detect a case based on symptoms^20–25^. It also accounts for the epidemiological and immunological characteristics of the Delta and Omicron variants^26^ and stratifies individuals by vaccination status. Specifically, we distinguished between unvaccinated individuals, those fully vaccinated with a primary vaccination course, and boosted individuals^27^. Vaccination coverage data were dynamically implemented and sourced from national registries^27^. Furthermore, we considered age-specific vaccine effectiveness against infection, transmission, and clinical symptoms given infection, depending on the number of doses, time since last dose, and the variant of concern^28–31^. Full details on the model structure, parameter values, and estimates are reported in the Supplementary Material (Supplementary Material, pp 4-12).

Sensitivity analyses were performed on the values of children susceptibility and transmissibility, and the residency time in the *R*_+_ compartment (Supplementary Material, pp 15-17)

### Simulations

We simulated the school protocols that were implemented in France in the study period (**Table 1**).

**Table 1.** Timeline of application of the school protocols in France in the study period (from week 45, 2021 to week 06, 2022).

| Study period | Period 1 – Delta wave (2021) |  |  |  |  |  | Period 2 – Omicron wave (2022) |  |  |  |  |  |
| --- | --- | --- | --- | --- | --- | --- | --- | --- | --- | --- | --- | --- |
| Week (2021-2022) | W45 | W46 | W47 | W48 | W49 | W50 | W01 | W02 | W03 | W04 | W05 | W06 |
| Nationwide protocols | Reactive class closure<br>Isolation: 7 days for confirmed symptomatic cases<br>Quarantine: 10 days for all the classmates<br>Test type: Saliva samples analyzed by RT-PCR<br>Turnaround time : 1 day |  |  |  | Reactive screening after 1 and 7 days from the symptomatic case detection<br>Isolation: 10 days for all positive cases<br>Quarantine : 7 days for all the classmates whenever three confirmed cases appeared in a class, 10 days for not compliant students<br>Test type: Saliva samples analyzed by RT-PCR or LDF<br>Turnaroun d time : 1 day |  | Strengthened reactive screening at d0, after 2, and after 4 days from the symptomatic case detection<br>Isolation: 7 days for all positive cases<br>Test type: Saliva samples analyzed by RT-PCR or LFD<br>Turnaround time : 1 day |  |  |  |  |  |
| Experimental protocols | / |  | Weekly screening<br>Isolation: 7 days for all positive cases<br>Quarantine: 7 days for all the classmates whenever three confirmed cases appeared in a class<br>Test type: Salivary Saliva samples analyzed by RT-PCR<br>Turnaround time : 1 day |  |  |  |  |  |  |  |  |  |

The model fit was performed on prevalence estimates derived from the experimental weekly screening protocol, starting in week 47, 2021. Simulations started in week 45, when schools reopened after a two-week break, allowing us to assume that transmission chains within schools were interrupted. During weeks 45 and 46 of 2021, we simulated the reactive class closure protocol implemented in France at the time, which required a seven-day class closure following the detection of a positive case. Starting in week 47, this was replaced by the weekly screening protocol, which mandated a seven-day isolation for students testing positive and a class closure if three or more cases were detected. Separate fits were performed for the Delta wave (Period 1) and Omicron wave (Period 2). Age-specific seroprevalence estimates and vaccination coverage were used to initialize the model^27,32^. Weekly introductions were stochastically estimated using age-specific community surveillance data by department and adjusted to account for detection rate and estimated within-school transmission dynamics^10^.

All simulated protocols assumed the use of salivary PCR tests with a one-day turnaround time. Test sensitivity was modeled as age-dependent and time-varying, with a peak sensitivity of 96%^33^ (Supplementary Material, p. 11).

To assess the effectiveness of the weekly screening protocol in reducing infections, we compared it numerically to protocols implemented in other schools of the same departments during the same period. We computed averted school transmission in the weeks when weekly screening was applied (i.e., weeks 47-50 of 2021 in Period 1 and weeks 01-06 of 2022 in Period 2). Specifically, we parameterized the model with the transmissibility β*MLE* and weekly introductions estimated under the weekly screening protocol. Using these parameters, we simulated the reactive class closure protocol applied between weeks 45 and 48 of 2021, followed by the reactive screening protocol starting in week 49. Following the changes in protocol, in Period 1 the simulated reactive screening required tests on days 1 and 7 for students in the same class as a positive case, with a ten-day isolation period for positive cases and class closure if three cases were detected in a week. In Period 2, we simulated the strengthened reactive screening protocol, which required tests on days 0, 2, and 4 for students in the same class as a positive case, with a seven-day isolation period but no class closure. For all counterfactual scenario simulations, the participation rate within each class was fixed at 68%, corresponding to the median adherence observed in the experimental weekly screening.

### Inference framework

We used a maximum likelihood estimation (MLE) approach to fit the model to the student prevalence observed in primary schools implementing the experimental weekly screening. We estimated the transmissibility per contact per unit time (β/min) at school for the Delta and Omicron variants, considering time-varying adherence rates based on the observed participation data from each department.

In Period 1, the detection rate in the community was treated as a free parameter. It was explored in a grid and selected using the Akaike Information Criterion (AIC) score to optimize model fit. In Period 2, the detection rate was estimated by comparing age-specific community surveillance data for children aged 5–11 years with the prevalence reported in schools.

Transmissibility estimates for both periods were obtained from 2,000 simulated stochastic outbreaks per parameter set over a 6-week timeframe (42 days). Sensitivity analyses were conducted to examine the effects of cohorting and the impact of reduced test sensitivity for asymptomatic individuals during the pre-symptomatic and post-symptomatic phases.

### Comparison with prior estimates from the Alpha wave

We extended the analyses we performed in a previous work to study SARS-CoV-2 transmission in primary schools during the Alpha wave^10^ to estimate the contribution of within-school transmission in that wave and compare it with the results of this study from the Delta and Omicron waves. Details are available in the Supplementary Material, p. 15.

## RESULTS

A total of 28,643 tests were performed as part of the experimental weekly screening protocol in schools in the Auvergne-Rhône-Alpes region (**Figure 1**). During Period 1, 14,630 tests were collected in 24 schools across four departments: 3,103 tests in 5 schools in Isère, 1,230 in 2 schools in Puy-de-Dôme, 7,434 in 9 schools in Rhône, and 2,863 in 8 schools in Savoie. In Period 2, 14,013 tests were carried out in 14 schools: 550 tests in 3 schools in Isère, 13,143 in 10 schools in Rhône, and 320 in 1 school in Savoie. After applying the inclusion criteria, the total number of tests was reduced to 10,297 in Period 1 (7,434 tests in 9 schools in Rhône and 2,863 tests in 8 schools in Savoie) and 13,143 in Period 2, all of which were from the Rhône department where 10 schools decided to continue adopting the experimental protocol beyond the initial study period (**Table 1**). For sensitivity, we conducted the analysis on all schools participating to the experimental protocol, resulting in additional 5 primary schools in the Isère department and 2 primary schools in Puy-de-Dôme in Period 1 (see Supplementary Material pp. 4, 13).

Adherence rates varied across departments and periods. In Period 1, Rhône showed the highest adherence, ranging from 70% to 82% with an increasing trend over time (**Table 2**). In Savoie, adherence increased from 59% to 75% before dropping to 68% by the end of the period. In Period 2, adherence in Rhône rose in parallel with the Omicron surge, peaking during week 03 of 2022.

**Table 2.** Number of tests, students at school, observed adherence, and prevalence by weeks and departments in the primary schools participating in the experimental weekly screening and included in the study.

|  |  | Period 1 – Delta wave (2021) |  |  |  | Period 2 – Omicron wave (2022) |  |  |  |  |  |
| --- | --- | --- | --- | --- | --- | --- | --- | --- | --- | --- | --- |
|  |  | W47 | W48 | W49 | W50 | W01 | W02 | W03 | W04 | W05 | W06 |
| **Tests** | Rhône | 1369 | 1847 | 2111 | 2107 | 1769 | 2246 | 2514 | 2503 | 1827 | 2284 |
|  | Savoie | 537 | 692 | 804 | 830 | / | / | / | / | / | / |
| **Students** | Rhône | 1965 | 2713 | 2590 | 2566 | 2482 | 3021 | 3006 | 3057 | 3070 | 3072 |
|  | Savoie | 912 | 1063 | 1066 | 1213 | / | / | / | / | / | / |
| **Adherence** | Rhône | 69.7 | 68.1 | 81.5 | 82.1 | 71.3 | 74.3 | 83.6 | 81.9 | 59.5 | 74.3 |
|  | Savoie | 58.9 | 65.1 | 75.4 | 68.4 | / | / | / | / | / | / |
| **Prevalence** | Rhône | 1.53 | 2.33 | 1.33 | 0.95 | 5.03 | 8.06 | 8.15 | 5.07 | 5.09 | 3.20 |
|  | Savoie | 1.30 | 2.17 | 2.24 | 0.96 | / | / | / | / | / | / |

During the first two weeks following the school holidays (weeks 45 and 46), participating schools adhered to the reactive class closure protocol, which was applied nationwide, before the experimental weekly screening was implemented. In Period 1, prevalence rates in participating schools peaked during the second week of the protocol (week 48) in Rhône and the third week (week 49) in Savoie (**Figure 2**). In Period 2, the prevalence peak in Rhône occurred during the third week after schools reopened. **Table 2** summarizes the number of tests, number of students, adherence rates, and prevalence after applying the inclusion criteria. The corresponding information on the full dataset are reported in the Supplementary Material (Supplementary Material, pp 2-4).

**Figure 2.**
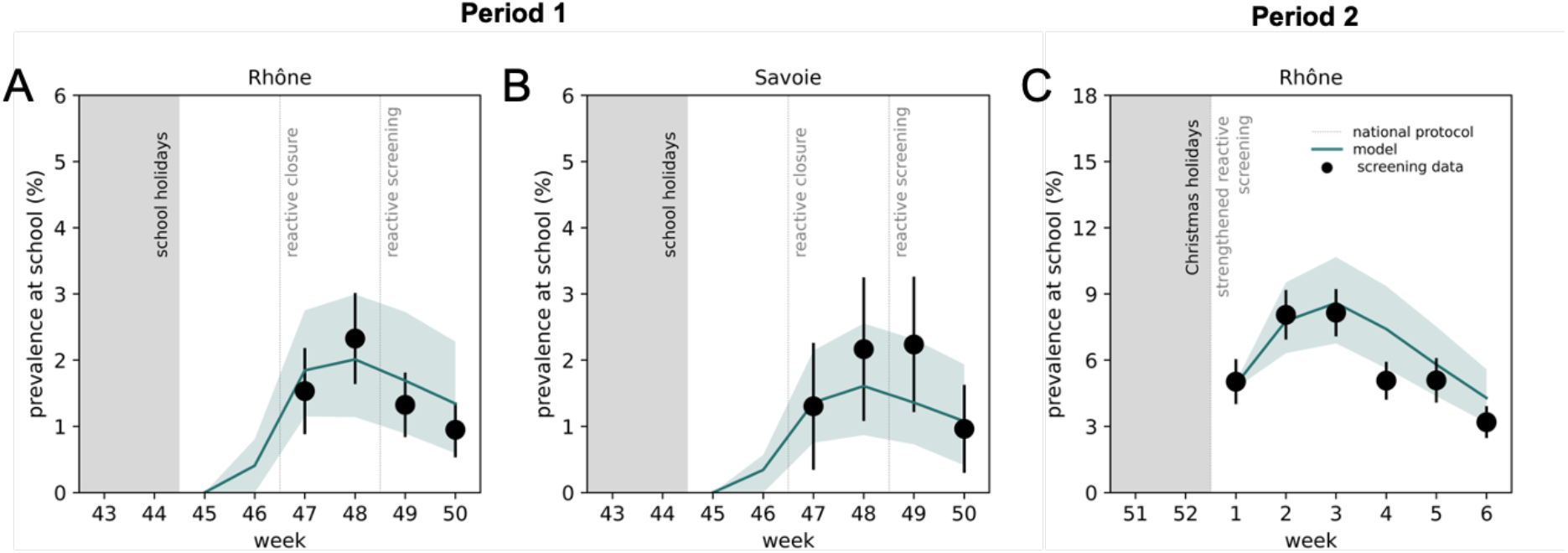
Estimated and observed prevalence at school in Period 1 and Period 2 of experimental weekly screening in the Rhône and Savoie departments. **A**.Green line represents the prevalence estimated by the model in Period 1 of experimentation during the Delta wave in the Rhône department selected through the inclusion criteria. The green area represents the interquartile range (IQR). The black dots represent the observed prevalence at school as obtained by experimental screening data. The error bars represent 95% confidence intervals. The grey area corresponds to holiday periods. Dashed grey lines show the protocols in place at the national level **B**. As in **A** for the Savoie department in Period 1 during the Delta wave. **C**. As in **A** for the Rhône department in Period 2 during the Omicron period. In panels **1A** and **1B**, the simulated school protocols were reactive class closure from week 45 to week 46, 2021 and experimental weekly screening from week 47 to week 50 of 2021. In panel **1C**, the simulated school protocol was the weekly screening from week 01 to week 06 of 2022. In all the panels, we considered salivary PCR tests.

The prevalence predicted by the fitted model closely matched the observed data for the departments included in the study: Rhône and Savoie during Period 1 (**Figure 2A, 2C**) and Rhône during Period 2 (**Figure 2B**). To evaluate the contribution of within-school transmission, we compared the fit estimates for the number of infections acquired at school and those introduced from the community (Supplementary Material, p 14). In Period 1, within-school transmission accounted for 67% of all infections in both departments with IQR ranges of 53–78% for Rhône and 50–82% for Savoie (**Figure 3A**). In Period 2, within-school transmission represented 52% (IQR 47– 57%) of total infections (**Figure 3B**). Sensitivity analyses confirmed the stability of these estimates when varying model parameters, including extended durations in the post-symptomatic compartment (*R*_+_) (10 days instead of 6), reduced test sensitivity for asymptomatic and pre-symptomatic phases, and variations in cohorting effects (Supplementary Material, p 21). Estimates of the contribution of within-school transmission during the Delta and Omicron waves were similar to those obtained during the Alpha wave in the departments of the same region (64% (IQR 20%-81%) in the Ain department; 62% (IQR 14%-80%) in the Loire department; 59% (IQR 17%-76%) in Rhône department, see Supplementary Material p. 15).

**Figure 3.**
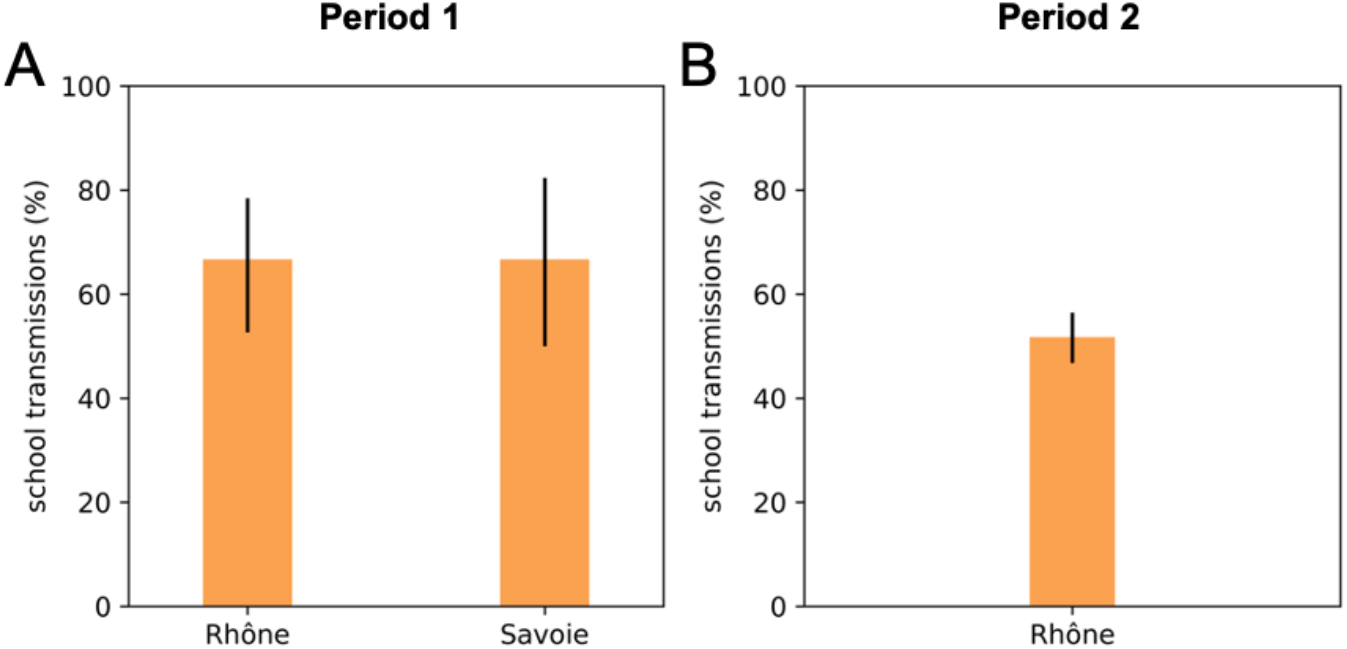
Estimated school transmission contribution in Period 1 and Period 2 of experimental weekly screening under the Delta and Omicron waves by departments. **A**.Bars show the school transmission contribution predicted by the model in the selected departments of Rhône and Savoie in Period 1 of the experimentation (i.e., weeks 47-50 of 2021) during the Delta wave. The error bars represent the interquartile range (IQR) **B**. As in A Period 2, corresponding to the Omicron wave.

To compare the experimental weekly screening to the nationally implemented reactive strategies, we modelled school prevalence assuming the same number of introductions. During Period 1, the nationally applied reactive screening strategy, introduced in week 49 of 2021, was predicted to result in peak student prevalence rates of 3.0% (IQR 1.9–4.6%) in Rhône and 2.1% (IQR 1.1–3.5%) in Savoie. These values were 50% and 30% higher, respectively, than the peaks observed under weekly screening (**Figure 4A, 4C**). Additionally, school prevalence peaks under the reactive strategies occurred later compared to weekly screening, which allowed for earlier curve flattening. In Period 2, the strengthened reactive screening strategy applied between weeks 01 and 06 of 2022 was predicted to result in a school peak prevalence of 11.7% (IQR 9.4-14.1) in Rhône, which was 40% higher than the peak observed under the experimental weekly screening (**Figure 4B**).

**Figure 4.**
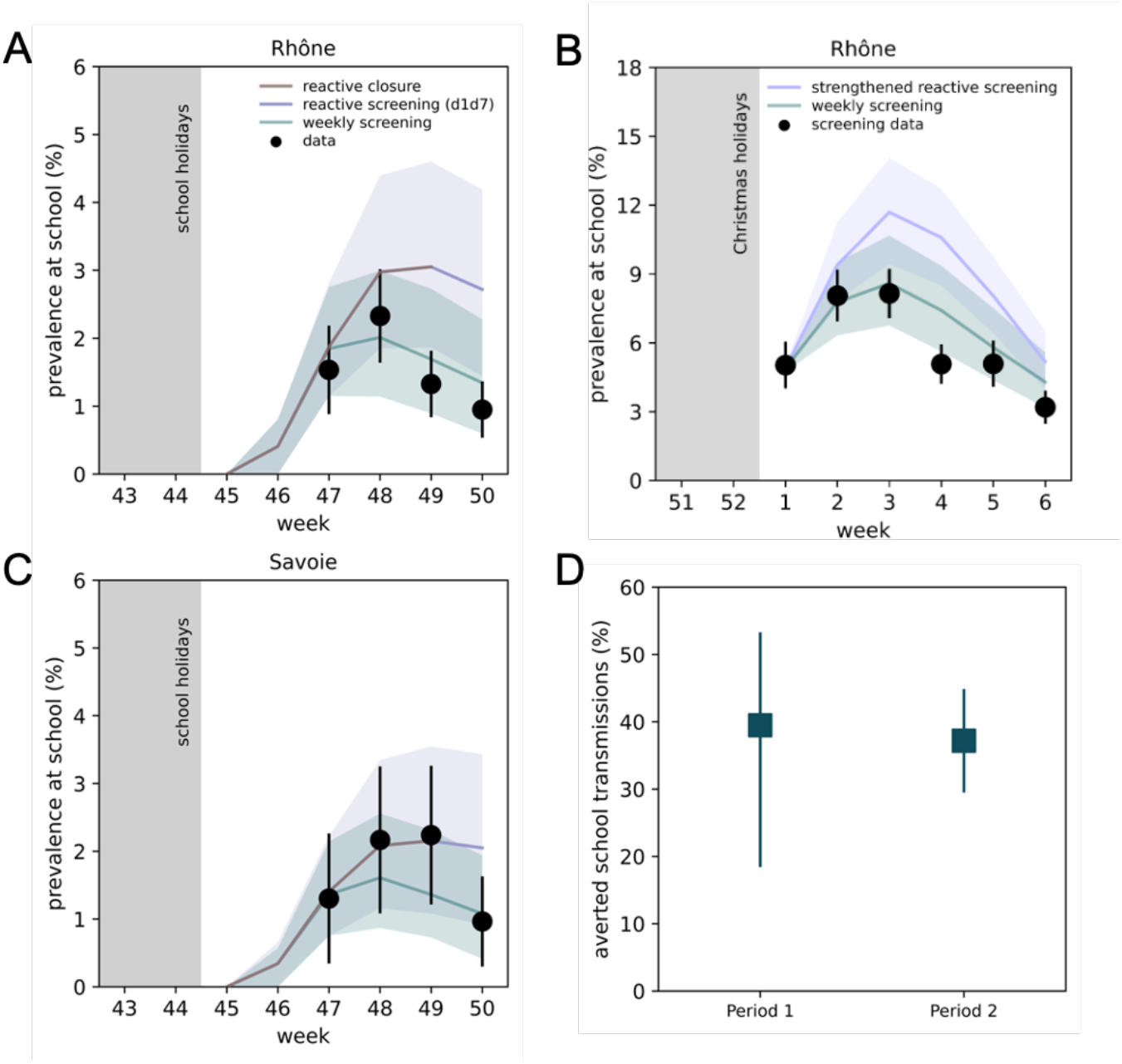
Comparison between estimated prevalence under the experimental weekly screening and the national reactive strategies by departments and estimated averted school transmissions. **A**.Lines represent the prevalence estimated by the model in the Rhône department across various school protocols: reactive class closure from week 45 to week 48 (brown line), reactive screening on days 1 and 7 from week 49 to week 50 (violet line), and experimental weekly screening from week 47 to week 50 of 2021 (green line). Areas represent the interquartile ranges (IQR). The black dots represent the observed prevalence at school as obtained by experimental screening data. The error bars represent 95% confidence intervals. **B** As in panel A, but for Period 2 (week 01 to week 06, 2022) during the Omicron wave, when strengthened national reactive screening on days 0, 2, and 4 was implemented at the national. **C**. As in A for Savoie department. **D**. Percentage of averted school transmissions achieved by weekly screening compared to the reactive strategies in the two periods of experimentation (i.e., weeks 47-50 of 2021 in Period 1 and weeks01-06 of 2022 in Period 2). In all the panels, we considered salivary PCR tests.

Weekly screening was estimated to avert 40% (IQR 18–53%) of within-school transmissions during Period 1 and 37% (IQR 30–45%) during Period 2 compared to the reactive strategies (**Figure 4D**). These estimates remained consistent across sensitivity analyses, even when varying key parameters (Supplementary Material, pp. 21–23).

## DISCUSSION

We analysed virological test results from an experimental weekly screening protocol conducted in 17 primary schools in the Rhône and Savoie departments in France during the Delta wave (November 22–December 17, 2021), and in 10 primary schools in the Rhône department during the Omicron wave (January 3–February 13, 2022). We estimated that between half and two-thirds of student infections originated from school contacts, with systematic weekly screening reducing school-related transmissions by approximately 40% compared to the national reactive strategies across both periods.

A key advantage of weekly screening was its ability to identify and isolate cases earlier than reactive protocols, effectively reducing student prevalence. Notably, prevalence began to decline one week earlier under the weekly screening protocol compared to predictions for reactive screening. This earlier decline highlights the efficacy of systematic screening in mitigating viral spread by promptly identifying and isolating pre-symptomatic and asymptomatic cases, which account for a substantial portion of SARS-CoV-2 transmission, particularly in children^21,34,35^. These findings align with prior studies in England showing that twice-weekly testing in England successfully controlled within-school transmission in secondary schools^36^, and reinforce numerical evidence on the benefits of regular screening^12,37–41^.

Participation rates were critical to the success of the protocol. In Period 1, adherence reached over 80% in Rhône and over 67% in Savoie by the end of the period, contributing significantly to the flattening of the prevalence curve. Similarly, in Period 2, adherence in Rhône exceeded 80% during the Omicron peak (weeks 03–04 of 2022), coinciding with a reduction in student prevalence. These results are consistent with previous modeling studies that suggest high adherence—at least three-quarters of students— is essential for achieving meaningful case reduction^10,13^. However, such high adherence levels were not achieved during pilot screenings conducted in the same region during the Alpha wave, where adherence rates hovered around 50% in primary schools and as low as 10% in middle and high schools^10^. Sustaining such high adherence requires well-coordinated, effective communication strategies that address community concerns and encourage participation^42,43^. For example, authorities in Baselland, Switzerland, successfully implemented weekly salivary PCR testing with adherence rates exceeding 75%^13^ before making participation mandatory in 2022.

Proactive screening proved more effective than reactive screening strategies in detecting asymptomatic and pre-symptomatic cases. Children, who are less likely to develop symptoms than adults^21^, often remain undetected under reactive screening protocols, allowing infections to spread silently. Supporting this, the COVID-19 Schools Infection Survey in England (2020–2021) found that most cases identified through screening were asymptomatic^44^, while a screening campaign in Piedmont, Italy, during early 2021 revealed that asymptomatic cases accounted for a substantial portion of infections during periods of high community transmission^45^. These findings, together with prior modeling studies^11,12,41,46^, underscore the importance of proactive screening in addressing asymptomatic transmission and reducing school-related infections.

In contrast, reactive screening was ineffective in controlling infections and presented significant operational challenges. During the study period, the continuous occurrence of COVID-19 cases triggered surges in test demand^13^, overburdening testing centers and straining pharmacies and laboratories. The lack of predictability associated with reactive screening, which depends on the detection of symptomatic cases, further complicated resource allocation and planning. This unpredictability causes additional stress for educators and parents, particularly during the Omicron wave. Teachers in France expressed concerns about the continuity of education ^47^, while families struggled to access timely testing, often resulting in children staying home unnecessarily and parents missing work.

Proactive screening, while resource-intensive, offers a more organized and structured alternative, reducing logistical uncertainties and minimizing disruptions. However, implementing such programs remains challenging, particularly in low- and middle-income countries. According to the second UNESCO-UNICEF-World Bank survey on COVID-19 school responses (May–August 2020), only 19% of countries reported plans for school-based testing, and only 50% of low- and middle-income countries reported sufficient resources to implement even basic health protocols^48^. Bridging these gaps requires strategic investments in education and health infrastructure, ensuring that proactive interventions are accessible and tailored to the unique needs of local contexts^49^. Such efforts are critical for achieving equitable responses to future health crises.

The contribution of schools to SARS-CoV-2 transmission varied across periods and contexts. In this study, we estimated that 50-70% of infections detected through weekly screening originated in schools, with a lower proportion during the Omicron wave likely due to increased community transmission during the Christmas break when schools were closed. These estimates are consistent with findings from the Alpha wave in the same region. Based on prevalence data from 71 primary schools in Ain, Loire, and Rhône^10^, we estimated that schools contributed 59% to 64% of infections during the five weeks preceding the third lockdown implemented to curb the Alpha wave (weeks 8-13, 2021). Comparable results were reported in Belgium, where reconstruction of SARS-CoV-2 outbreaks in a primary school using epidemiological and genomic data found that 66% of transmissions originated in schools, with most transmission occurring between children or from children to adults^50^. Similarly, an analysis of 87 school outbreaks in Italy during March–April 2021 attributed nearly half of confirmed cases among school attendees to school contacts under reactive class closure protocols^51^. Our findings differ from those of a modeling study on U.S. primary schools participating in a weekly testing program from Spring 2021 to March 2022, which estimated a lower contribution of schools to overall infections^52^. These differences may stem from the use of a compartmental model, which did not explicitly account for detailed heterogeneous interaction patterns, or from differences in the broader epidemiological and policy context.

Additional studies from the US^53^, Sweden^54^, and France^55^ also identified higher infection risks in households with school-going children or teachers attending in person. These results emphasize that while schools are not the primary amplification hubs for SARS-CoV-2 as they are for influenza^56^, they do play a significant role in transmission, particularly in the absence of robust mitigation strategies. Secondary attack rates in educational settings remained relatively low when non-pharmaceutical interventions (NPIs) such as masking and ventilation were implemented. For instance, studies from Switzerland^57^ and Italy^58^ demonstrated that mask mandates and mechanical ventilation significantly reduced transmission risks. Future research should further investigate the role of environmental factors like ventilation in controlling transmission, as well as how household spillover from school infections contributes to community spread^59,60^.

Our study has a set of limitations. First, we did not explicitly assess aerosol transmission or the effectiveness of ventilation measures during the experimental period. 93.1% of schools reported applying natural ventilation more than twice daily, alongside compulsory mask use during Period 1. However, our estimation of the transmission rate inherently accounts for the implemented preventive measures. Second, our analysis was limited to school-based transmission and did not account for infections introduced into households. Prior research suggests that active school surveillance can reduce such spillover effects^60^. Third, our findings are limited to primary schools and may not generalize to secondary schools, where student density, mixing patterns, and activities – as well as student epidemiological and immunological characteristics – differ. Further research in real-world conditions is needed to assess effectiveness of weekly screening in these contexts. Our previous modeling work showed that weekly screening remains the most effective strategy in controlling viral spread in high schools compared to reactive protocols^10^, highlighting its broader potential across educational levels. Additionally, while systematic screening is resource-intensive, its testing demand under high-incidence conditions is comparable to reactive strategies requiring three tests over four days^13^. Even with reduced diagnostic sensitivity, such as with antigen tests, systematic screening was shown to consistently outperform reactive approaches in controlling disease spread^10,13^. This underscores its robustness as an effective strategy for managing school-based transmission across varied contexts.

The COVID-19 pandemic pushed authorities worldwide to repeatedly close schools, exposing students to significant learning losses, increased anxiety, missed social opportunities, and exacerbated inequalities^61,62^. These consequences demonstrate that prolonged school closures are unsustainable, even during public health crises^63^. Flexible strategies that adapt to evolving transmission risks should be considered to optimize the balance between public health measures and educational needs. Systematic screening offers a viable and proactive strategy to mitigate pre-symptomatic and asymptomatic transmission, ensuring safer access to education while highly transmissible variants circulate. Our findings provide evidence to improve the design of non-pharmaceutical interventions in schools and contribute to future pandemic preparedness plans to make schools more resilient in the event of a future pandemic.

## Data Availability

De-identified individual data on contacts of the primary school under study are publicly available at the SocioPatterns project website. De-identified aggregated COVID-19 community surveillance data by age class are publicly available at SpF data observatory platform. De-identified aggregated COVID-19 prevalence data from experimental weekly screenings used in this study are contained in the manuscript.

http://www.sociopatterns.org/datasets/

## AUTHORS’ CONTRIBUTIONS

V.C. conceived and designed the study. B.L., C.E., P.V. carried out the experimental screening strategies at school and collected the data. E.C. analyzed the data, developed the code, and ran the simulations. All authors interpreted the results. E.C. and V.C. wrote the initial manuscript draft. All authors edited and approved the final version of the Article.

## DATA SHARING STATEMENT

De-identified individual data on contacts of the primary school under study are publicly available at the SocioPatterns project website (http://www.sociopatterns.org/datasets/). De-identified aggregated COVID-19 community surveillance data by age class are publicly available at Santé publique France data observatory platform (https://geodes.santepubliquefrance.fr/). De-identified aggregated COVID-19 prevalence data from experimental weekly screenings during the Delta and Omicron waves used in this study are available in the tables reported in the manuscript and Supplementary Material.

## ACKNOWLEDGMENTS

This study was partly funded by Agence Nationale de la Recherche project DATAREDUX (ANR-19-CE46-0008-03), EU Horizon 2020 grant MOOD (H2020-874850; paper catalogued as MOOD 125), Horizon Europe grants VERDI (101045989) and ESCAPE (101095619). The contents of this publication are the sole responsibility of the authors and don’t necessarily reflect the views of the European Commission.

## References

1. Littlecotta, H. et al. Measures implemented in the school setting to contain the COVID-19 pandemic. Cochrane Database of Systematic Reviews (2024) doi:10.1002/14651858.CD015029.pub2.

2. UNESCO. Education: From disruption to recovery. UNESCO Building peace in the minds of men and women https://en.unesco.org/covid19/educationresponse (2020).

3. National Education Responses to COVID-19 Summary report of UNESCO’s online survey.pdf.

4. The Impact of School Closures on Learning and Mental Health of Children: Lessons From the COVID-19 Pandemic - Deni Mazrekaj, Kristof De Witte, 2024. https://journals.sagepub.com/doi/10.1177/17456916231181108?utm_source=chatgpt.com.

5. Patrinos, H. A., Vegas, E. & Carter-Rau, R. An Analysis of COVID-19 Student Learning Loss.

6. Protocole sanitaire applicable dans les écoles et établissements scolaires à partir du 29 novembre 2021. IH2EF https://www.ih2ef.gouv.fr/protocole-sanitaire-applicable-dans-les-ecoles-et-etablissements-scolaires-partir-du-29-novembre.

7. Bénet, T. et al. Évaluation d’une campagne de dépistages itératifs hebdomadaires du SARS-CoV-2 en écoles maternelles et élémentaires : une étude prospective multicentrique. Médecine et Maladies Infectieuses Formation 2, S60 (2023).

8. Le protocole sanitaire de l’année scolaire 2021-2022. gouvernement.fr http://www.gouvernement.fr/actualite/le-protocole-sanitaire-de-l-annee-scolaire-2021-2022.

9. Santé publique France. COVID-19 : point épidémiologique du 25 novembre 2021. Santé publique France https://www.santepubliquefrance.fr/maladies-et-traumatismes/maladies-et-infections-respiratoires/infection-a-coronavirus/documents/bulletin-national/covid-19-point-epidemiologique-du-25-novembre-2021 (2021).

10. Colosi, E. et al. Screening and vaccination against COVID-19 to minimise school closure: a modelling study. The Lancet Infectious Diseases 22, 977–989 (2022).

11. McGee, R. S., Homburger, J. R., Williams, H. E., Bergstrom, C. T. & Zhou, A. Y. Model-driven mitigation measures for reopening schools during the COVID-19 pandemic. PNAS 118, (2021).

12. Torneri, A. et al. Controlling SARS-CoV-2 in schools using repetitive testing strategies. eLife 11, e75593 (2022).

13. Colosi, E. et al. Minimising school disruption under high incidence conditions due to the Omicron variant in France, Switzerland, Italy, in January 2022. Eurosurveillance 28, 2200192 (2023).

14. Santé publique France. Le point épidémiologique - Surveillance épidémiologique en région Auvergne-Rhône-Alpes. (2022).

15. Le protocole sanitaire en milieu scolaire simplifié. Gouvernement.fr https://www.gouvernement.fr/actualite/le-protocole-sanitaire-en-milieu-scolaire-simplifie.

16. Stehlé, J. et al. High-Resolution Measurements of Face-to-Face Contact Patterns in a Primary School. PLOS ONE 6, e23176 (2011).

17. Hart, W. S. et al. Generation time of the alpha and delta SARS-CoV-2 variants: an epidemiological analysis. The Lancet Infectious Diseases 22, 603–610 (2022).

18. He, X. et al. Temporal dynamics in viral shedding and transmissibility of COVID-19. Nature Medicine 26, 672–675 (2020).

19. Young, B. E. et al. Epidemiologic Features and Clinical Course of Patients Infected With SARS-CoV-2 in Singapore. JAMA 323, 1488–1494 (2020).

20. Goldstein, E., Lipsitch, M. & Cevik, M. On the Effect of Age on the Transmission of SARS-CoV-2 in Households, Schools, and the Community. J Infect Dis 223, 362–369 (2021).

21. Davies, N. G. et al. Age-dependent effects in the transmission and control of COVID-19 epidemics. Nature Medicine 26, 1205–1211 (2020).

22. Fontanet, A. et al. SARS-CoV-2 infection in schools in a northern French city: a retrospective serological cohort study in an area of high transmission, France, January to April 2020. Eurosurveillance 26, 2001695 (2021).

23. Dattner, I. et al. The role of children in the spread of COVID-19: Using household data from Bnei Brak, Israel, to estimate the relative susceptibility and infectivity of children. PLOS Computational Biology 17, (2021).

24. Han, M. S. et al. Clinical Characteristics and Viral RNA Detection in Children With Coronavirus Disease 2019 in the Republic of Korea. JAMA Pediatrics 175, 73 (2021).

25. Smith, L. E. et al. Adherence to the test, trace, and isolate system in the UK: results from 37 nationally representative surveys. BMJ 372, n608 (2021).

26. Powell, A. A. et al. Protection against symptomatic infection with delta (B.1.617.2) and omicron (B.1.1.529) BA.1 and BA.2 SARS-CoV-2 variants after previous infection and vaccination in adolescents in England, August, 2021–March, 2022: a national, observational, test-negative, case-control study. The Lancet Infectious Diseases 0, (2022).

27. Géodes - Santé publique France - Indicateurs : cartes, données et graphiques. https://geodes.santepubliquefrance.fr/#view=map2&c=indicator.

28. Dorabawila, V. et al. Effectiveness of the BNT162b2 vaccine among children 5-11 and 12-17 years in New York after the Emergence of the Omicron Variant. 2022.02.25.22271454 Preprint at 10.1101/2022.02.25.22271454 (2022).

29. Cohen-Stavi, C. J. et al. BNT162b2 Vaccine Effectiveness against Omicron in Children 5 to 11 Years of Age. N Engl J Med NEJMoa2205011 (2022) doi:10.1056/NEJMoa2205011.

30. Sacco, C. et al. Effectiveness of BNT162b2 vaccine against SARS-CoV-2 infection and severe COVID-19 in children aged 5–11 years in Italy: a retrospective analysis of January–April, 2022. The Lancet 400, 97–103 (2022).

31. Eyre, D. W. et al. Effect of Covid-19 Vaccination on Transmission of Alpha and Delta Variants. New England Journal of Medicine (2022) doi:10.1056/NEJMoa2116597.

32. Santé publique France. Séroprévalence Du SARS-CoV-2 En Population Générale, France Entière : Résultats Préliminaires de La Semaine 42 de 2021 (18-23 Octobre). (2021).

33. Smith, R. L. et al. Longitudinal Assessment of Diagnostic Test Performance Over the Course of Acute SARS-CoV-2 Infection. The Journal of Infectious Diseases 224, 976–982 (2021).

34. Dong, Y. et al. Epidemiology of COVID-19 Among Children in China. Pediatrics 145, e20200702 (2020).

35. Shekerdemian, L. S. et al. Characteristics and Outcomes of Children With Coronavirus Disease 2019 (COVID-19) Infection Admitted to US and Canadian Pediatric Intensive Care Units. JAMA Pediatr 174, 868 (2020).

36. Leng, T. et al. Quantifying pupil-to-pupil SARS-CoV-2 transmission and the impact of lateral flow testing in English secondary schools. Nat Commun 13, 1106 (2022).

37. Paltiel, A. D., Zheng, A. & Walensky, R. P. Assessment of SARS-CoV-2 Screening Strategies to Permit the Safe Reopening of College Campuses in the United States. JAMA Netw Open 3, e2016818 (2020).

38. Asgary, A., Cojocaru, M. G., Najafabadi, M. M. & Wu, J. Simulating preventative testing of SARS-CoV-2 in schools: policy implications. BMC Public Health 21, 125 (2021).

39. Bilinski, A. et al. Estimated Transmission Outcomes and Costs of SARS-CoV-2 Diagnostic Testing, Screening, and Surveillance Strategies Among a Simulated Population of Primary School Students. JAMA Pediatrics 176, 679–689 (2022).

40. Lopman, B. et al. A modeling study to inform screening and testing interventions for the control of SARS-CoV-2 on university campuses. Sci Rep 11, 5900 (2021).

41. Leng, T. et al. Quantifying within-school SARS-CoV-2 transmission and the impact of lateral flow testing in secondary schools in England. medRxiv preprint 2021.07.09.21260271 (2021) doi:10.1101/2021.07.09.21260271.

42. Watson, D. et al. How do we engage people in testing for COVID-19? A rapid qualitative evaluation of a testing programme in schools, GP surgeries and a university. BMC Public Health 22, 305 (2022).

43. Denford, S. et al. Feasibility and acceptability of daily testing at school as an alternative to self-isolation following close contact with a confirmed case of COVID-19: a qualitative analysis. BMC Public Health 22, 742 (2022).

44. COVID-19 Schools Infection Survey Round 4, England - Office for National Statistics. https://www.ons.gov.uk/peoplepopulationandcommunity/healthandsocialcare/conditionsanddiseases/bulletins/covid19schoolsinfectionsurveyround4england/antibodydatamarch2021.

45. Bena, E. F., Ilenia Eboli, Teresa Spadea, Carlo Saugo, Lorenzo Richiardi, Milena Maule, Pietro Presti, Antonella. ‘Scuola sicura’: a school screening testing programme to prevent the spread of COVID-19 in students in Piedmont. https://www.epiprev.it https://epiprev.it/articoli_scientifici/scuola-sicura-a-school-screening-testing-programme-to-prevent-the-spread-of-covid-19-in-students-in-piedmont.

46. Lasser, J. et al. Assessing the impact of SARS-CoV-2 prevention measures in Austrian schools using agent-based simulations and cluster tracing data. Nat Commun 13, 554 (2022).

47. Fermetures de classes, enseignants touchés, grèves… Pourquoi le variant Omicron risque de chahuter la rentrée scolaire. Franceinfo https://www.francetvinfo.fr/sante/maladie/coronavirus/variant-omicron/fermetures-de-classes-enseignants-touches-greves-pourquoi-le-variant-omicron-risque-de-chahuter-la-rentree-scolaire_4900807.html (2022).

48. UNESCO, UNICEF, & World Bank. What Have We Learnt?: Overview of Findings from a Survey of Ministries of Education on National Responses to COVID-19. (Paris, New York, Washington D.C.: UNESCO, UNICEF, World Bank, 2020). doi:10.1596/34700.

49. Jourdan, D. & Gray, N. J. Keeping schools open in times of health crises: seeing the global picture. The Lancet Child & Adolescent Health 8, 249–250 (2024).

50. Kremer, C. et al. Reconstruction of SARS-CoV-2 outbreaks in a primary school using epidemiological and genomic data. Epidemics 44, 100701 (2023).

51. Grané, C. M. et al. SARS-CoV-2 transmission patterns in educational settings during the Alpha wave in Reggio-Emilia, Italy. Epidemics 100712 (2023) doi:10.1016/j.epidem.2023.100712.

52. Huang, C. & Smith, R. L. A modeling study on SARS-CoV-2 transmissions in primary and middle schools in Illinois. BMC Public Health 24, 3197 (2024).

53. Lessler, J. et al. Household COVID-19 risk and in-person schooling. Science 372, 1092–1097 (2021).

54. Vlachos, J., Hertegård, E. & B.Svaleryd, H. The effects of school closures on SARS-CoV-2 among parents and teachers. Proceedings of the National Academy of Sciences 118, e2020834118 (2021).

55. Galmiche, S. et al. Exposures associated with SARS-CoV-2 infection in France: A nationwide online case-control study. The Lancet Regional Health - Europe 7, (2021).

56. Neil-Sztramko, S. E. et al. What is the specific role of schools and daycares in COVID-19 transmission? A final report from a living rapid review. The Lancet Child & Adolescent Health 8, 290–300 (2024).

57. Banholzer, N. et al. SARS-CoV-2 transmission with and without mask wearing or air cleaners in schools in Switzerland: A modeling study of epidemiological, environmental, and molecular data. PLOS Medicine 20, e1004226 (2023).

58. Buonanno, G., Ricolfi, L., Morawska, L. & Stabile, L. Increasing ventilation reduces SARS-CoV-2 airborne transmission in schools: A retrospective cohort study in Italy’s Marche region. Frontiers in Public Health 10, (2022).

59. Garcia-Bernardo, J., Hedde-von Westernhagen, C., Emery, T. & van Hoek, A. J. Assessing COVID-19 transmission through school and family networks using population-level registry data from the Netherlands. Sci Rep 14, 31248 (2024).

60. Galmiche, S. et al. Risk of SARS-CoV-2 Infection Among Households With Children in France, 2020-2022. JAMA Network Open 6, e2334084 (2023).

61. Theberath, M. et al. Effects of COVID-19 pandemic on mental health of children and adolescents: A systematic review of survey studies. SAGE Open Med 10, 20503121221086712 (2022).

62. The State of the Global Education Crisis: A Path to Recovery. World Bank https://www.worldbank.org/en/topic/education/publication/the-state-of-the-global-education-crisis-a-path-to-recovery.

63. Conducting after-action reviews of the public health response to COVID-19: update. https://www.ecdc.europa.eu/en/publications-data/conducting-after-action-reviews-public-health-response-covid-19-update-0 (2023).

